# Specific diagnostic method for St. Louis Encephalitis Virus using a non-structural protein as antigen

**DOI:** 10.1101/19002030

**Authors:** M.B. Simari, S.E. Goñi, V.C Luppo, C.M. Fabbri, M.H. Argüelles, M.E. Lozano, M.A. Morales, N.G. Iglesias

## Abstract

St. Louis encephalitis virus (SLEV) is a mosquito-borne reemerging flavivirus in Argentina. It is currently necessary to develop specific serological tests that can efficiently discriminate the flaviviruses that circulate in our country. The immunoassays to diagnose SLEV lack specificity because they are based on the detection of structural viral proteins and the human immunoglobulins produced during infection against these proteins cross-react with other flaviviruses. Here, we describe an enzyme-immunoassay designed to detect human IgG antibodies specific to the viral nonstructural protein NS5. The results indicate that NS5 is a promising antigen useful to discriminate SLEV from other circulating flaviviruses.

## Manuscript

St. Louis encephalitis virus (SLEV) is a *Flavivirus* (*Flaviviridae* family) transmitted by *Culex spp*. mosquitoes to birds, humans and other vertebrates are incidental hosts(1). In Argentina, SLEV is a re-emerging arbovirus broadly distributed with the potential to give rise to epidemics of human encephalitis. The symptoms of infection in humans ranged from a simple febrile headache to meningoencephalitis, and the severity of the symptoms increases with the patient age (1-5). Cross-reactivity in diagnostic assays between SLEV and other flaviviruses complicates differential diagnosis(6). This is particularly challenging in Argentina where there are dengue virus (DENV) outbreaks frequently, history of jungle yellow fever, and Zika virus (ZIKV) activity was demonstrated and YFV vaccination is common in some regions (7-10). The primary antigen used in assays intended for IgM and IgG detection during flavivirus infections has been the viral envelope (E) structural protein. However, the similarity of the flavivirus E proteins causes problems with cross-reactivity and false-positive test results, particularly in the acute and early convalescent phases. Plaque reduction neutralization testing (PRNT) is the gold standard for serological diagnosis of flavivirus, but this test is expensive, labor-intensive, and requires live virus manipulation.

Regarding the specific antiviral response pattern against SLEV, IgM antibodies persistence has been detected in some patients showing that the presence of IgM might not be indicative of acute infection (11, 12). Spinsanti *et al*. have shown that in 57% of the samples studied IgM antibodies persisted after a year of the infection. However, in IgM positive samples the diagnosis should be confirmed by PRNT to differentiate among *Flavivirus*. They have also demonstrated the persistence of the four subclasses of IgG antibodies for more than a year and the presence of IgG early after infection(11).

SLEV is an icosahedral single-stranded RNA virus of approximately 11 kb that encodes a single polyprotein that is processed co- and post-translationally to produce three structural proteins and seven non-structural proteins involved in virus replication(13). The non-structural protein NS5 is the viral methyl-transferase and RNA-dependent RNA polymerase(13). NS5 is an essential protein for viral RNA synthesis with a mean similarity among flaviviruses of 68%.

Due to the high cross-reactivity of the E protein among flavivirus it has been proposed the use of non-structural proteins as antigens for diagnosis. It has been demonstrated that IgM and IgG antibodies against non-structural proteins NS1, NS2B, NS3, and NS5 are produced during flavivirus infections (14-20). Besides, antibodies against the non-structural proteins NS3 and NS5 have been detected in serum samples of patients infected with WNV and DENV(15, 16, 18-20), and an NS5-based assay has been shown to discriminate between WNV and DENV or SLEV infections(19). Recently, the use of recombinant NS5 of DENV serotype 2 as antigen has allowed differentiating serum samples of patients with primary and secondary DENV infections from serum samples of patients with JEV and WNV infections taken during the acute and convalescent phases(16, 18).

In this work, we developed an indirect enzyme immunoassay (EIA) using the purified full-length recombinant SLEV NS5 as antigen for the detection of IgG antibodies in human serum samples.

For antigen preparation, we cloned, expressed in *E. coli* and purified full-length NS5 from SLEV CbaAr-4005 strain. CbaAr-4005 is a genotype III strain of SLEV isolated from *Culex quinquefasciatus* mosquitoes in Córdoba, Argentina, in 2005, during the largest SLEV outbreak ever reported in South America(21). After 2005 outbreak, additional SLEV outbreaks in Argentina occurred in provinces of Entre Ríos (2006), Buenos Aires (2010), and San Juan(2011) (5).

The NS5 expression and purification were carried out as previously described for DENV NS5 (22). Briefly, the DNA fragment containing the coding region of full-length NS5 was PCR amplified and cloned into the plasmid pET-28at *Bam*HI and *Xho*I sites, adding a six-histidine tag at the amino terminus of the proteins. The protein was expressed in *E. coli* overnight at 20°C after induction with IPTG. The lysis of the cells was carried out using a French press in phosphate binding buffer in the presence of DNase I and protease inhibitor cocktail (Sigma). After centrifugation, NS5 was purified on a His-Trap nickel Sepharose affinity column (GE Healthcare) using the standard procedure. The protein was dialyzed against 20 mM Tris-HCl buffer, pH 7.5, 300 mM NaCl, 10% glycerol, and 1 mM dithiothreitol. The dialyzed protein solution was further purified by size exclusion chromatography using a Superdex 200 column. Figure 1 shows an SDS-PAGE of samples collected through the protein purification protocol. Approximately 3 mg of pure SLEV NS5 protein per liter of cell culture were obtained.

Anonymized barcode serum samples obtained from patients with febrile symptoms and flavivirus infection confirmed or discharged at the National Reference Laboratory (INEVH-ANLIS) were selected to evaluate the performance of the immunologic assay described in this article. Plaque-reduction neutralization test (PRNT) was used as the reference method for this evaluation. To perform this method, serum samples were heat inactivated at 56°C for 30 min; and two-fold serially diluted, from 1:20 to 1:2560. Replicates of all dilutions of each serum sample were incubated, at 37 °C for 1 h, with 100 plaque-forming units (PFU) of WNV (ChimeriVax TM WNV strain), SLEV (ChimeriVax TM SLEV strain), DENV-1 (Hawaii strain), DENV-2 (NGC strain), DENV-3 (H87 strain), DENV-4 (H241 strain), WNV(ChimeriVax TM WNV strain), YFV (vaccine 17D-YEL strain) and ZIKV(ARCB116141 strain). After incubation, 100 µl of each virus-serum mixture were inoculated onto a 12-well plate monolayer of Vero C76 cells (80% confluent growth) and absorbed for 1 h at 37°C. First overlay medium (containing low-melting point agarose (GIBCO BRL, Life Technologies) Hank’s BSS, vitamins, amino acids, heat-inactivated calf serum, L-glutamine, and 7.5% sodium bicarbonate) was added, 0.5 mL per well and allowed to solidify for 15 minutes. The plates where SLEV and WNV were tested were incubated for four days; and plates where DENV 1-4, YFV and ZIKV were assayed were incubated for 5 days. Neutral red staining was performed by adding a neutral red second overlay (same as above without fetal calf serum and containing 4% Neutral Red [Sigma Chemical, St. Louis, MO]), 0.5 mL per well and allowed to solidify for 15 minutes. Plates were incubated overnight at 37°C with 4% CO_2_ and plaques were counted and titers were calculated and expressed as the reciprocal of the serum dilution yielding a ≥90% reduction in PFU (PRNT90). The results of the PRNT are shown in table 1. Fifty-nine sera were analyzed, 19 were positive for SLEV, 14 DENV positive (85.7% DENV-1, 7.1 % DENV-2, 7.1 % without serotype), 9 YFV positive, 4 ZIKV positive, and 14 were negative for the analyzed viruses. Monotypic or heterotypic patterns were differentiated according to whether the serum was positive to one or several flaviviruses, respectively. In heterotypic patterns, interpretation of PRNT data was as follows: samples with a neutralizing antibody titer (PRNT90) ≥ four-fold higher than the other flavivirus titers were considered positive for antibody to that virus. Using these samples representative of the flavivirus that co-circulate in our country we evaluated our enzyme immunoassay. We developed a standard indirect immunoassay to detect SLEV NS5-specific IgG in human sera. For this purpose, we determined an optimum NS5 concentration of 500 ng/well and a dilution 1:100 of serum samples. The antigen was coated overnight at 4°C in a 96-microwell plate in a solution containing 50 mM Na_2_CO_3_, pH 9.6. After coating, the plates were washed three times with PBS and blocked with casein 0.2% in PBS (blocking solution) at 37°C for 1 h. Later, sera from patients were diluted 1:100 in blocking solution and incubated at 37°C for 2 h. Then, the plates were PBS washed and incubated with HRP conjugated anti-human IgG antibody (Abcam) 1:2000 at 37°C for 2h. After washing the plates, ABTS (KPL labs) peroxidase substrate was added and after 15 min the reaction was stopped with SDS and the optical density at 405 nm was determined.

**Table 1.** PRNT_90_ titers for 8 flaviviruses.

| ID | Days after symptoms onset | PRNT<sub>90</sub> Titer |  |  |  |  |  |  |  | Result Interpretation |
| --- | --- | --- | --- | --- | --- | --- | --- | --- | --- | --- |
|  |  | DENV 1 | DENV 2 | DENV 3 | DENV 4 | SLEV | WNV | YFV | ZIK V |  |
| 1 | 6 | <10 | <10 | <10 | <10 | 320 | 10 | 80 | <10 | SLEV |
| 2 | 8 | <10 | <10 | <10 | <10 | 320 | 10 | 80 | <10 | SLEV |
| 3 | 13 | <20 | <20 | <20 | <20 | 160 | <20 | <20 | <20 | SLEV |
| 4 | 25 | <20 | <20 | <20 | <20 | 80 | <20 | <20 | <20 | SLEV |
| 5 | 12 | <10 | 20 | 10 | <10 | 40 | 10 | <10 | <10 | SLEV |
| 6 | 57 | <10 | 20 | 10 | 10 | >80 | 10 | <10 | <10 | SLEV |
| 7 | 5 | <10 | <10 | <10 | <10 | 40 | <10 | <10 | <10 | SLEV |
| 8 | 24 | <10 | <10 | <10 | <10 | 160 | <10 | <10 | <10 | SLEV |
| 9 | 4 | <20 | <20 | <20 | <20 | 20 | <20 | <20 | <20 | SLEV |
| 10 | 16 | <40 | <40 | <40 | <40 | 40 | <40 | <40 | <40 | SLEV |
| 11 | 175 | <40 | <40 | <40 | <40 | 80/320 | <40 | <40 | <40 | SLEV |
| 12 | 5 | <20 | <20 | <20 | <20 | 40 | <20 | <20 | <20 | SLEV |
| 13 | 62 | <40 | <40 | <40 | <40 | 80 | <40 | <40 | <40 | SLEV |
| 14 | 30 | <40 | <40 | <40 | <40 | 80 | <40 | <40 | <40 | SLEV |
| 15 | 70 | <40 | <40 | <40 | <40 | 80 | <40 | 40 | <40 | SLEV |
| 16 | 8 | <40 | <40 | <40 | <40 | <40 | <40 | <40 | <40 | NEG |
| 17 | 8 | <20 | <20 | <20 | <20 | <20 | <20 | <20 | <20 | NEG |
| 18 | 9 | 160 | <20 | <20 | <20 | <20 | <20 | <20 | <20 | DENV-1 |
| 19 | 24 | 320 | 80 | 80 | 40 | <40 | <40 | <40 | <40 | DENV-1 |
| 20 | 13 | <40 | <40 | <40 | <40 | <40 | <40 | <40 | <40 | NEG |
| 21 | 25 | <20 | <20 | <20 | <20 | <20 | <20 | <20 | <20 | NEG |
| 22 | 1 | <20 | <20 | <20 | <20 | <20 | <20 | <20 | <20 | NEG |
| 23 | 15 | <20 | <20 | <20 | <20 | <20 | <20 | <20 | <20 | NEG |
| 24 | 7 | <20 | <20 | <20 | <20 | <20 | <20 | <20 | <20 | NEG |
| 25 | 20 | <40 | <40 | <40 | <40 | <40 | <40 | <40 | <40 | NEG |
| 26 | NA | <20 | <20 | <20 | <20 | <20 | <20 | <20 | <20 | NEG |
| 27 | 5 | <20 | <20 | <20 | <20 | <20 | <20 | <20 | 40 | ZIKV |
| 28 | 12 | <40 | <40 | <40 | <40 | <40 | <40 | <40 | 40 | ZIKV |
| 29 | 11 | 20 | 20 | <20 | <20 | <20 | <20 | <20 | <20 | DENV |
| 30 | 22 | <40 | 40 | <40 | <40 | <40 | <40 | <40 | <40 | DENV-2 |
| 31 | 9 | <20 | <20 | <20 | <20 | <20 | <20 | <20 | <20 | NEG |
| 32 | NA | <20 | <20 | <20 | <20 | <20 | <20 | 80 | <20 | YFV |
| 33 | NA | <20 | <20 | <20 | <20 | <20 | <20 | 20 | <20 | YFV |
| 34 | 12 | 80 | 20 | 20 | <20 | <20 | <20 | <20 | <20 | DENV-1 |
| 35 | 19 | 160 | <40 | <40 | <40 | <40 | <40 | <40 | <40 | DENV-1 |
| 36 | 17 | <40 | <40 | <40 | <40 | <40 | <40 | 320 | <40 | YFV |
| 37 | 32 | <20 | <20 | <20 | <20 | <20 | <20 | 160 | <20 | YFV |
| 38 | NA | <40 | <40 | <40 | <40 | <40 | <40 | >1280 | <40 | YFV |
| 39 | 8 | 160 | <20 | <20 | <20 | <20 | <20 | <20 | <20 | DENV-1 |
| 40 | 19 | 160 | 40 | 40 | <40 | <40 | <40 | <40 | <40 | DENV-1 |
| 41 | 8 | 320 | <20 | <20 | <20 | <20 | <20 | <20 | <20 | DENV-1 |
| 42 | 22 | >1280 | 40 | 40 | <40 | <40 | <40 | <40 | <40 | DENV-1 |
| 43 | 5 | <20 | <20 | <20 | <20 | 20 | <20 | <20 | <20 | SLEV |
| 44 | 69 | <40 | <40 | <40 | <40 | 40 | <40 | <40 | <40 | SLEV |
| 45 | 5 | <20 | <20 | <20 | <20 | <20 | <20 | 320 | <20 | YFV |
| 46 | NA | >640 | 20 | <20 | <20 | <20 | <20 | <20 | <20 | DENV-1 |
| 47 | NA | <40 | <40 | <40 | <40 | <40 | <40 | <40 | <40 | NEG |
| 48 | 10 | <20 | <20 | <20 | <20 | 20 | <20 | <20 | <20 | SLEV |
| 49 | 34 | <40 | <40 | <40 | <40 | 40 | <40 | <40 | <40 | SLEV |
| 50 | 16 | 160 | <20 | 20 | <20 | <20 | <20 | <20 | <20 | DENV-1 |
| 51 | 31 | 160 | <40 | <40 | <40 | <40 | <40 | <40 | <40 | DENV-1 |
| 52 | 8 | <20 | <20 | <20 | <20 | <20 | <20 | >640 | <20 | YFV |
| 53 | 18 | <40 | <40 | <40 | <40 | <40 | <40 | 640 | <40 | YFV |
| 54 | 22 | <20 | <20 | <20 | <20 | <20 | <20 | <20 | <20 | NEG |
| 55 | 7 | <40 | <40 | <40 | <40 | <40 | <40 | <40 | <40 | NEG |
| 56 | NA | 640 | 80 | 80 | <40 | <40 | <40 | <40 | <40 | DENV-1 |
| 57 | 17 | <40 | <40 | <40 | <40 | <40 | <40 | 160 | <40 | YFV |
| 58 | NA | <40 | <40 | <40 | <40 | 40 | 80 | <40 | 320 | ZIKV |
| 59 | NA | ND | ND | ND | ND | ND | ND | ND | ND | WNV* |

\*IgM e IgG positive in “in-house” ELISA.

**Figure 1.**
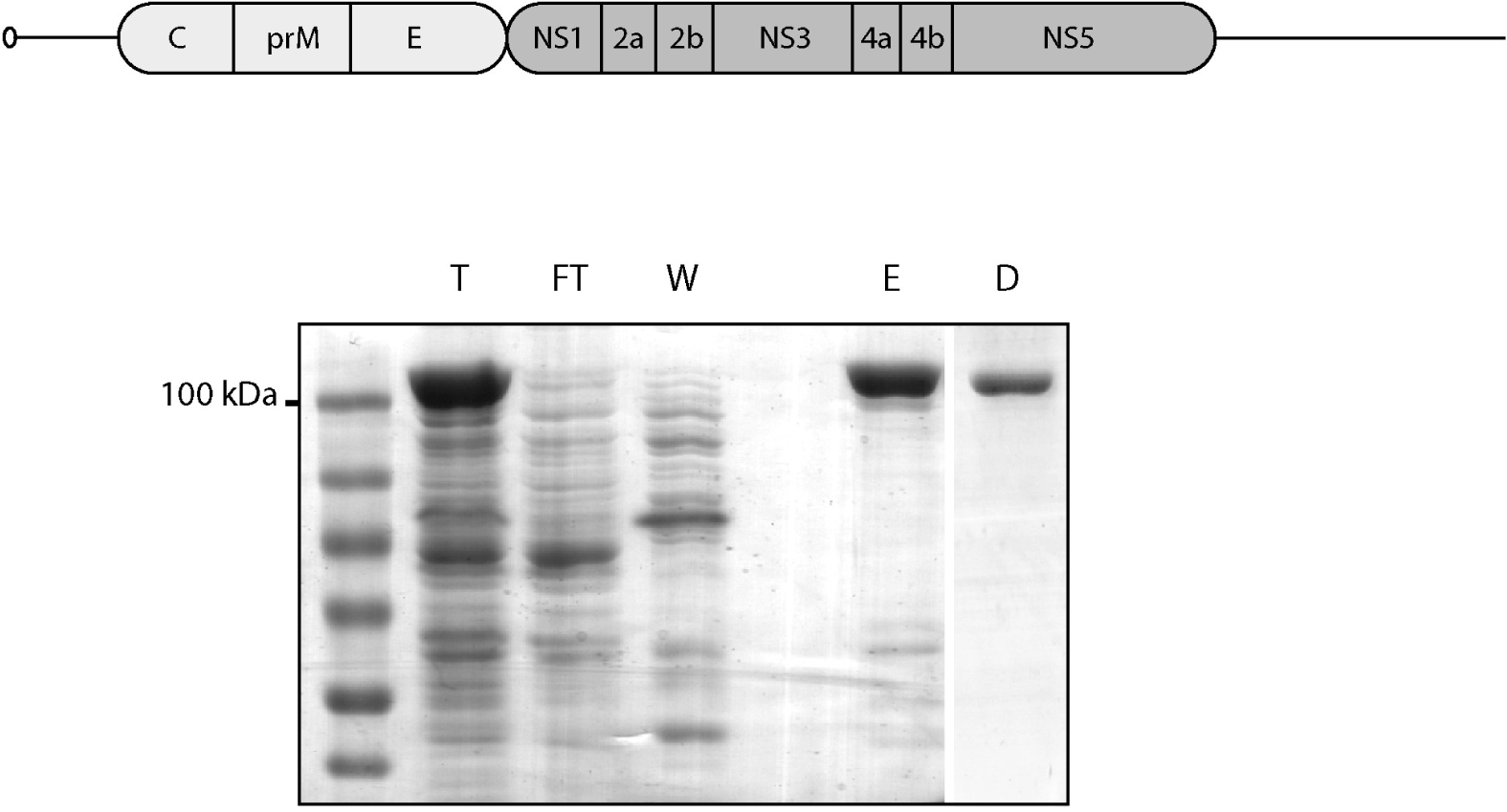
SLEV NS5 purification. At the upper panel SLEV genome structure. Lower panel coomassie stained SDS-PAGE: T, total soluble extract injected into IMAC column. FT, flow through. W, washed fraction. E, eluted protein from IMAC. D, Superdex purified and dialyzed protein.

Based on PRNT90 results we separated the samples into three groups: SLEV positive, other positive flavivirus and negative samples. The results clearly show that the assay efficiently discriminates SLEV positives samples from positive samples for other flavivirus or negative samples (Figure 2). Figure 3 shows the results for each individual serum sample. The cut-off to considerate a sample as positive by EIA was determined as 0.3, calculated as the mean value of negative samples plus two standard deviations. Taking into account this value, 16 out of 19 samples considered positive by PRNT were also positive by EIA (84%). Interestingly, the sera of SLEV infected patients (both homotypic and heterotypic pattern) can react with the recombinant NS5 protein, and no positive sample for DENV, YFV, ZIKV or WNV resulted positive in our assay. This is important since the presence of antibodies against NS5 in sera from patients infected with SLEV had never been reported. Using NS5 as an antigen for IgG detection we were able to avoid cross-reaction with antibodies to other flaviviruses. Although the number of serum positive samples for other flaviviruses was limited the results suggest that this test could be useful as a diagnostic method for SLEV infections. Further studies with a larger panel of serum samples will be necessary to validate this method. The three negative samples (9, 10, and 44) had low antibodies titers as judged by PRNT90, which could explain the negative result observed in our EIA. Although PRNT is the gold standard method we cannot rule out that the results observed for these samples are not due to cross reactivity against some other flavivirus not tested here (e.g., *Ilheus virus, Bussuquara virus*). The specificity of NS5-based EIA might reduce or eliminate the need for PRNT and therefore the requirement of BSL3 to the diagnosis of SLEV. Also, EIA is a 1 to 2-day assay compared with PRNT that is a 5 to 7-day assay.

In Argentina different flaviviruses that can produce similar clinical symptoms co-circulate. The laboratory diagnosis is crucial to determine the etiological agent causing the disease. Molecular methods (RT-PCR) are useful during the acute-phase, and then serological methods should be used. In our assay, we were able to detect IgG antibodies against NS5 in samples collected from five to 175 days after the onset of the symptoms (Table 1 and figure 3), this includes the acute and the convalescent phase.

**Figure 2.**
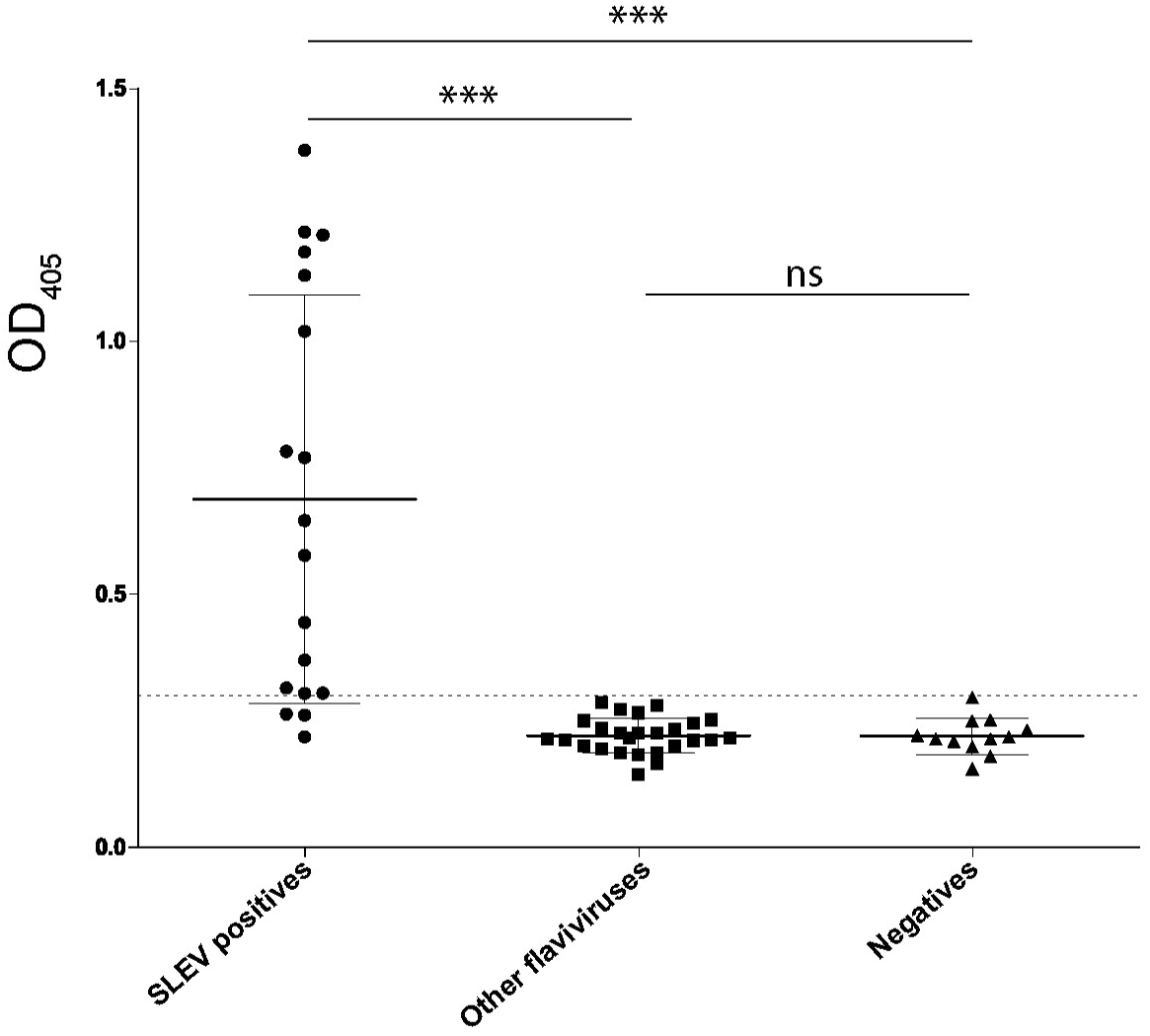
NS5 immuno-assay from serum samples. The error bars indicate the standard deviations. Statistical significance by one-way ANOVA analysis.

**Figure 3.**
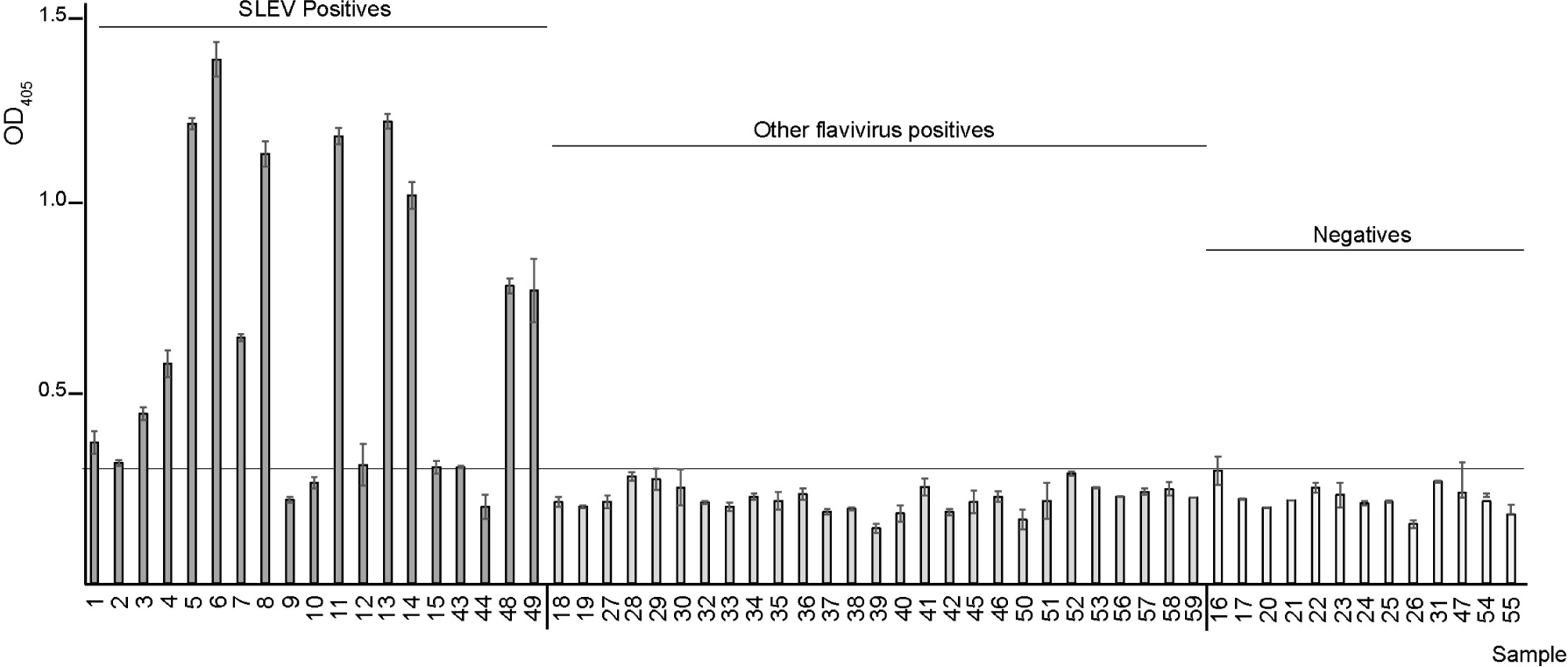
EIA NS5 IgG detection from individual samples. Results showed as mean ± SD (*** p<0.01, ns: non-significant).

The complexity of flavivirus diagnosis requires developing novel diagnostic methods, that, combined with those existing, allow us properly identifying these viruses.

In conclusion, we show that the EIA test based on NS5 is a potential diagnostic method for SLEV that avoids cross-reactivity with other flaviviruses and implies a less laborious and expensive method, safer and feasible to introduce in laboratories of low complexity.

## Data Availability

All data of this manuscript are available.

## Acknowledgements

This work was funded by grants from Universidad Nacional de Quilmes and Florencio Fiorini Foundation. NGI and MEL are members of the Argentinean Council of Investigation (CONICET). MBS was granted with a CIC-BA (Commission of Scientific Investigations of Buenos Aires, Argentina) fellowship. We are grateful to Laboratorio de Arenavirus y Arbovirus, Instituto de Virología “Dr. J.M. Vanella”, Facultad de Ciencias Médicas, Universidad Nacional de Córdoba (UNC), Argentina, for the genetic material of SLEV CbaAr-4005 strain.

***The authors declare that there is no conflict of interest regarding the publication of this article***.

## Notes

### Competing Interest Statement

The authors have declared no competing interest.

### Author Declarations

All relevant ethical guidelines have been followed and any necessary IRB and/or ethics committee approvals have been obtained.

Any clinical trials involved have been registered with an ICMJE-approved registry such as ClinicalTrials.gov and the trial ID is included in the manuscript.

